# Quantifying Care, Qualifying Experiences: A Systematic Review of Measurement-Based Care in Psychiatry from Patient and Provider Perspectives

**DOI:** 10.1101/2025.04.13.25323641

**Authors:** Ayan Dey, Ze’ev Lewis, Josh Posel, Rachel Pan, Karen Wang

## Abstract

**Objective:** This systematic review synthesizes clinician and patient perspectives on the benefits and drawbacks of measurement-based care (MBC) in psychiatry.

**Study Selection and Analysis:** We searched Ovid MEDLINE, EMBASE, EBM Reviews, APA PsychINFO, and CINAHL databases from inception to January 2024. After screening 1643 titles and abstracts, 47 full papers were reviewed, and 23 studies were ultimately included. Quality assessment was conducted using the Mixed Methods Appraisal Tool, and key patterns were extracted using thematic analysis.

**Findings:** The review reflects opinions of 657 patients and 2830 clinicians across various settings. Patients valued MBC for enhancing communication, self-awareness, and reducing stigma. However, they expressed concerns about the adequacy of measures in reflecting their clinical state and uncertainty about how responses influence treatment decisions. Clinicians appreciated MBC for improving patient involvement, tracking treatment response, and enhancing communication efficiency. Concerns included inadequate capture of clinical complexity, potential reporting biases, time constraints, insufficient training, and concerns with respect to data usage and privacy.

**Conclusions:** While patients and clinicians recognize significant benefits, including enhanced communication, improved insight, and more structured clinical decision-making, they also identify important limitations. These include concerns about the adequacy of scales to capture complex clinical presentations, potential impacts on the therapeutic alliance, and increased administrative burden. Moving forward, successful integration of MBC into routine care will require addressing these challenges through improved clinician training, clear guidelines for interpretation, greater transparency with respect to how data will be used, and more seamless integration with existing clinical workflows.

**Systematic Review Registration:** PROSPERO CRD420250651562

SUMMARY BOX

What is already known on this topic?
Measurement-based care (MBC) in psychiatry involves using validated scales to systematically evaluate patient symptoms and progress. Despite evidence supporting its potential to enhance treatment efficacy and patient engagement, implementation of MBC in clinical practice remains limited.

What this study adds?
This systematic review synthesizes perspectives from 657 patients and 2830 clinicians, gathered from 23 studies, on the use of MBC in psychiatric practice across various settings. It reveals shared appreciation for improved communication, treatment monitoring, and patient self-awareness. However, it also highlights concerns about the adequacy of measures in capturing clinical complexity, potential negative impacts on the therapeutic relationship, concerns with respect to use of data for performance monitoring, and implementation challenges.

How might this study affect research, practice or policy?
The findings emphasize the need for developing more comprehensive and adaptive assessment tools and improving clinician training in MBC implementation and interpretation. Future efforts should focus on increasing standardization, transparency, and simplification of MBC processes, including integration into electronic medical records systems. This could enhance mental health care delivery by addressing identified limitations while leveraging the benefits of increased patient engagement and treatment monitoring.

## 1. Introduction

Measurement-based care (MBC) describes the evidence-based practice of using validated questionnaires and scales to systematically evaluate patient progress on symptoms and functioning throughout treatment – helping to inform treatment decisions.[1] In response to growing demands for more patient-centered care, MBC has become increasingly common across healthcare systems.[2] This is particularly pronounced in psychiatry and clinical psychology, where validated scales aid in symptom tracking and improving standardization, while also facilitating efficient communication between providers.[2] When combined with clinical judgement, MBC enhances treatment efficacy by detecting poor response and subtle signs of clinical deterioration that may otherwise be missed in the early stages.[3] It can also improve treatment efficiency by helping clinicians efficiently identify problem areas for targeted symptom management.[1] MBC can also improve patient engagement by allowing more opportunity for shared decision making.[1, 4] For example, it can help patients become more mindful of their own progress and facilitate recognition of early signs of relapse or signs of improvement. This increased engagement in turn can facilitate improved adherence to medication and treatment protocols.

The rationale for this systematic review stems from the discrepancy between the empirical support for MBC and its limited adoption in clinical practice. Despite much evidence showing that MBC enhances mental health treatment efficacy, therapeutic efficiency, and patient adherence,[5] less than 20% of behavioural health practitioners integrate it into their practice.[6] This gap between research and implementation necessitates inquiry into barriers to its adoption. That said, this review aims to systematically examine existing literature on patient and physician perspectives towards MBC, identify the various ways clinicians currently use MBC in their practice, explore perceptions of MBC’s impact on patient experience and engagement, and investigate potential unintended negative effects of MBC implementation. By addressing these objectives, this study seeks to bridge the gap between the empirical support for MBC and its limited adoption in clinical practice, potentially informing strategies to increase its implementation in mental health care settings.

## 2. Methods

The study protocol is available at PROSPERO (CRD420250651562). Findings are presented according to the Preferred Reporting Items for Systematic Reviews and Meta-Analysis (PRISMA) guidelines.[7] Manuscript title was amended from the time that the protocol was registered. The PRISMA checklist can be found online in **supplemental index 1**.

### 2.1 Selection criteria

The published literature was searched for quantitative randomised controlled trials (RCTs), quasi-experimental studies, cross-sectional and pre–post study designs, qualitative and mixed-method studies. Case reports, commentaries and review articles were excluded studies. The eligibility criteria are provided in **Table 1**.

**Table 1.**
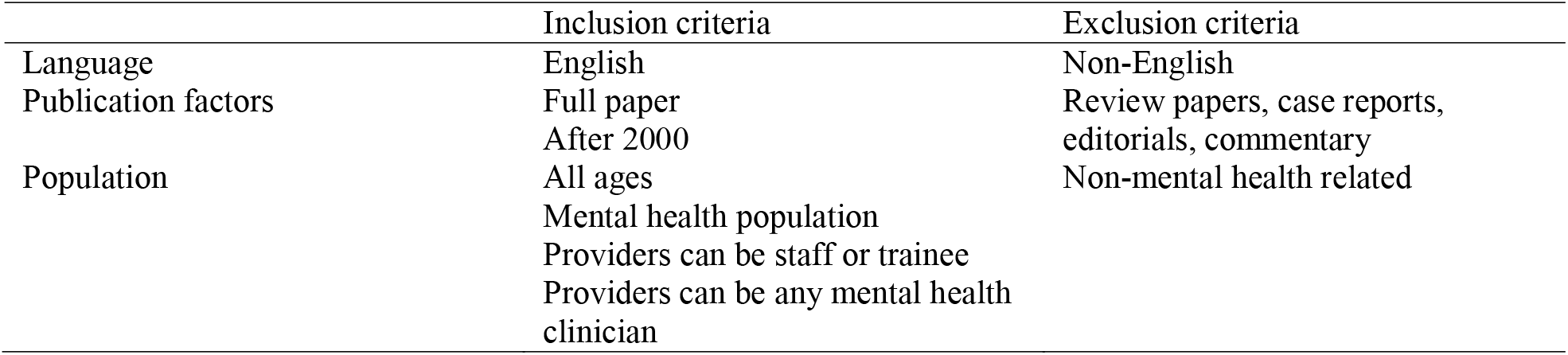
Inclusion and exclusion criteria.

### 2.2 Search Strategy

The search strategy was developed for the electronic databases to identify journal articles. The following databases were searched for related studies: Ovid MEDLINE (1946 to present), EMBASE (Classic EMBASE + EMBASE ((1947 to present)), EBM Reviews – Cochrane Central Register of Controlled Trials (1991 to January 2024), APA PsychINFO (1806 to June 2024), and CINAHL (via EBSCO ((1981 to current)). The search strategy was designed to comprehensively capture literature related to measurement-based care in mental health, with a focus on patient and physician perspectives. Specifically, the search combined three main concepts: 1) mental health including MeSH terms and keywords for psychiatry, mental disorders, anxiety disorders, bipolar disorder and mood disorders; 2) measurement based care, encompassing terms related to routine outcome monitoring, patient-reported measures, and feedback-informed treatment in mental health settings; and 3) patient and physician perspectives, incorporating MeSH terms and keywords to capture viewpoints and attitudes of both patients and healthcare providers. These concepts were combined using Boolean operators, with results limited to English language and human studies. Notably while, searches in the electronic databases were conducted from database inception until January 2024. The search was later limited to studies reported in English, involving human subjects. While initially there was no limit on publication year, only papers published during or after 2000 were included. The full search strategy is described in the online **supplemental index 2**.

### 2.3 Study selection

The search results were uploaded to the systematic review management software Covidence to remove duplicates. Two reviewers (AKD and KW) independently screened the remaining studies’ titles and abstracts against inclusion/exclusion criteria. Any uncertainty surrounding the inclusion of a study or disagreement was resolved through discussion between the reviewers – when in doubt papers were considered for full text review. Full texts were then screened against inclusion/exclusion criteria by reviewers (AKD, KW, JP, ZL, and RP), with each paper being independently reviewed by at least two reviewers. Inter-rater agreement between reviewers ranged from 0.77 to 1. Any uncertainty surrounding inclusion of a study was then resolved through discussion among the two reviewers. Any disagreement following discussions between the reviewers were resolved through discussion among all 5 reviewers and a majority vote.

### 2.4 Data extraction

All five reviewers (AKD, KW, JP, ZL and RP) independently extracted data from the included studies. The same reviewers who had completed the full text review also extracted the data. AKD reviewed all the data extraction tables. If there were any discrepancies during this sampling check, discussion took place and for final clarification a third, impartial reviewer was consulted. Data were extracted from published studies using a data extraction form in Microsoft Excel and included details of authors, study setting, study population, methodology, as well as descriptions of patient and provider perspectives (benefits and limitations) of measurement-based care.

### 2.5 Quality assessment

Quality assessment was done by same reviewers as the extraction using the Mixed Methods Appraisal Tool.[8] This was done in order to provide an overall description of the quality of studies included in this review. Descriptors “low”, “medium” and “high” were used to provide an indication of the quality of included studies. For quantitative, qualitative and cross-sectional studies, if a study met all five criteria it was classified as “high quality”. Whereas studies that met three to four criteria were classified as “medium quality” and studies that met only one or two criteria were classified as “low quality”. For mixed-method studies, the overall score was based on the lowest score of each of the study components (quantitative and qualitative). In the case of disagreements between reviewers, the final decision was taken after discussion with an independent third reviewer.

### 2.6 Data analysis

Given the diversity of settings, populations, the non-uniformity of interventions and the variations in outcome measures, a statistical meta-analysis was not appropriate. Hence, the review used a narrative synthesis. This approach allowed for a comprehensive and flexible analysis of the heterogeneous data extracted from the included studies. First, a preliminary synthesis was conducted to organize the findings from included studies and identify patterns across the data. The robustness of the synthesis was then assessed by considering the quality and relevance of the evidence, as well as any potential biases. Individual findings from studies were merged into themes through consensus between reviewers.

## 3. Results

### 3.1 Study selection

The findings of the search strategy are summarised in **Figure 1** as a Preferred Reporting Items for Systematic Reviews and Meta-Analyses (PRISMA) flow chart.[7] After removal of duplicates a total of 1418 titles and abstracts were screened. From these 47 full papers were reviewed and 23 papers were included in the final review. The references of included studies were reviewed for potential studies and no new studies were included. A total of 23 articles were excluded for the following reasons: unrelated to measurement-based care (n =14); not a full paper (n =4), editorial/commentary (n =2), non-mental health-related (n =1), and study focused on screening rather than patient/provider perspectives (2).

**Figure 1:**
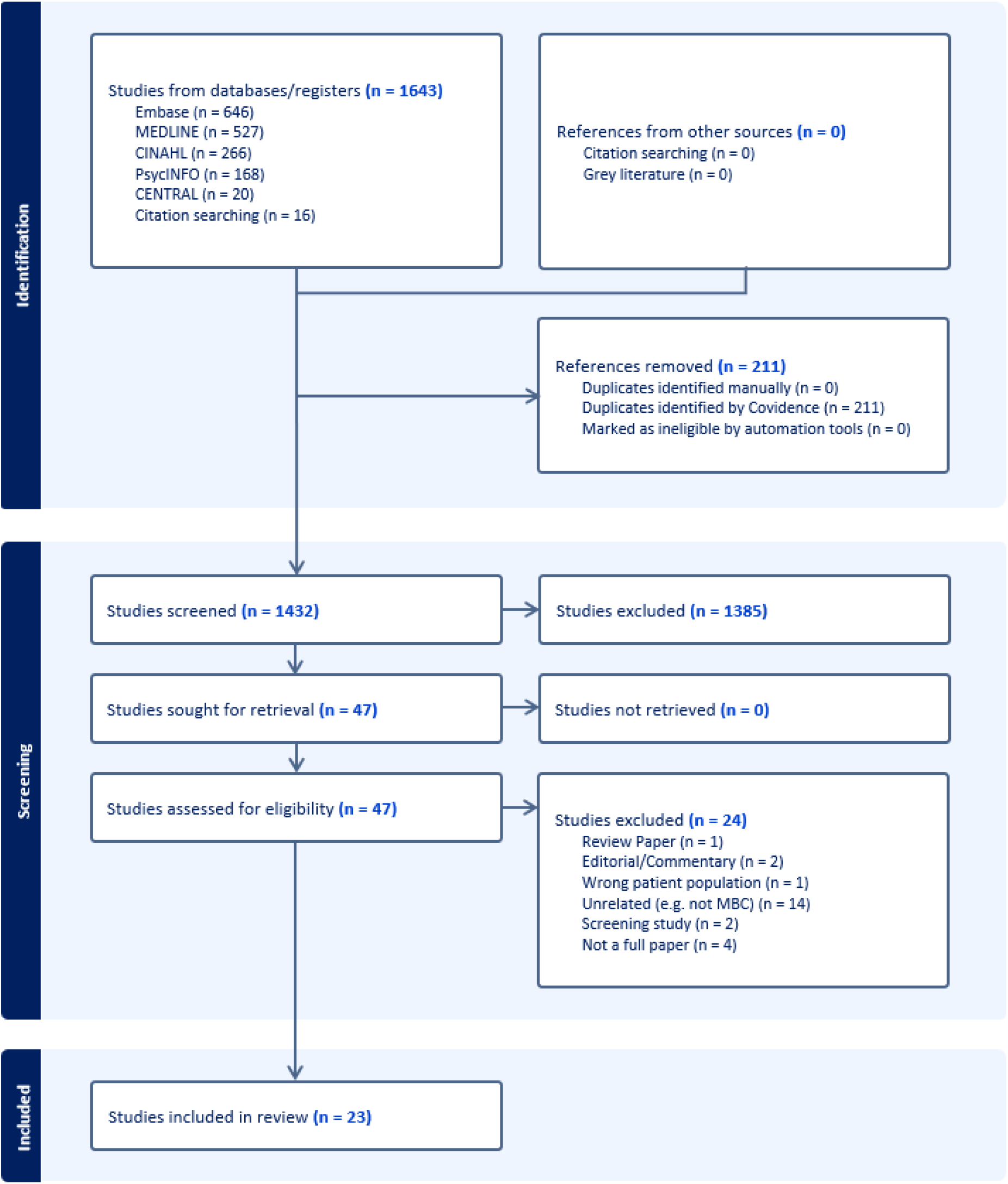
Preferred Reporting Items for Systematic Reviews and Meta-Analyses flow diagram[9]. MBC, measure-based care.

### 3.2 Characteristics of included studies

The designs of included studies were mixed methods (n = 5), observational (n = 8), and qualitative (n = 10). Cumulatively across studies this survey reflects the opinions and attitudes of a total of 901 service users/patients and 2814 clinicians across various professions (psychiatry, psychologists, therapists, and nurses). Practice settings also varied across studies; mixed (n = 1), outpatient (n = 19), and inpatient (n = 3). Characteristics of the included studies are reported in online **supplementary table S1**.

### 3.3 Quality appraisal

Of the 23 studies included, 12 were judged to be of “medium quality”, 9 were judged to be of “high quality” and only two were judged as “low quality” – indicating that this review primarily most reflects the results of well-designed studies. This is also reported in online **supplementary table S1**.

### 3.4 Synthesis of results from included studies

Several themes emerged from our review of the literature which are summarized in **Table 2** and described in the following sections.

**Table 2.**
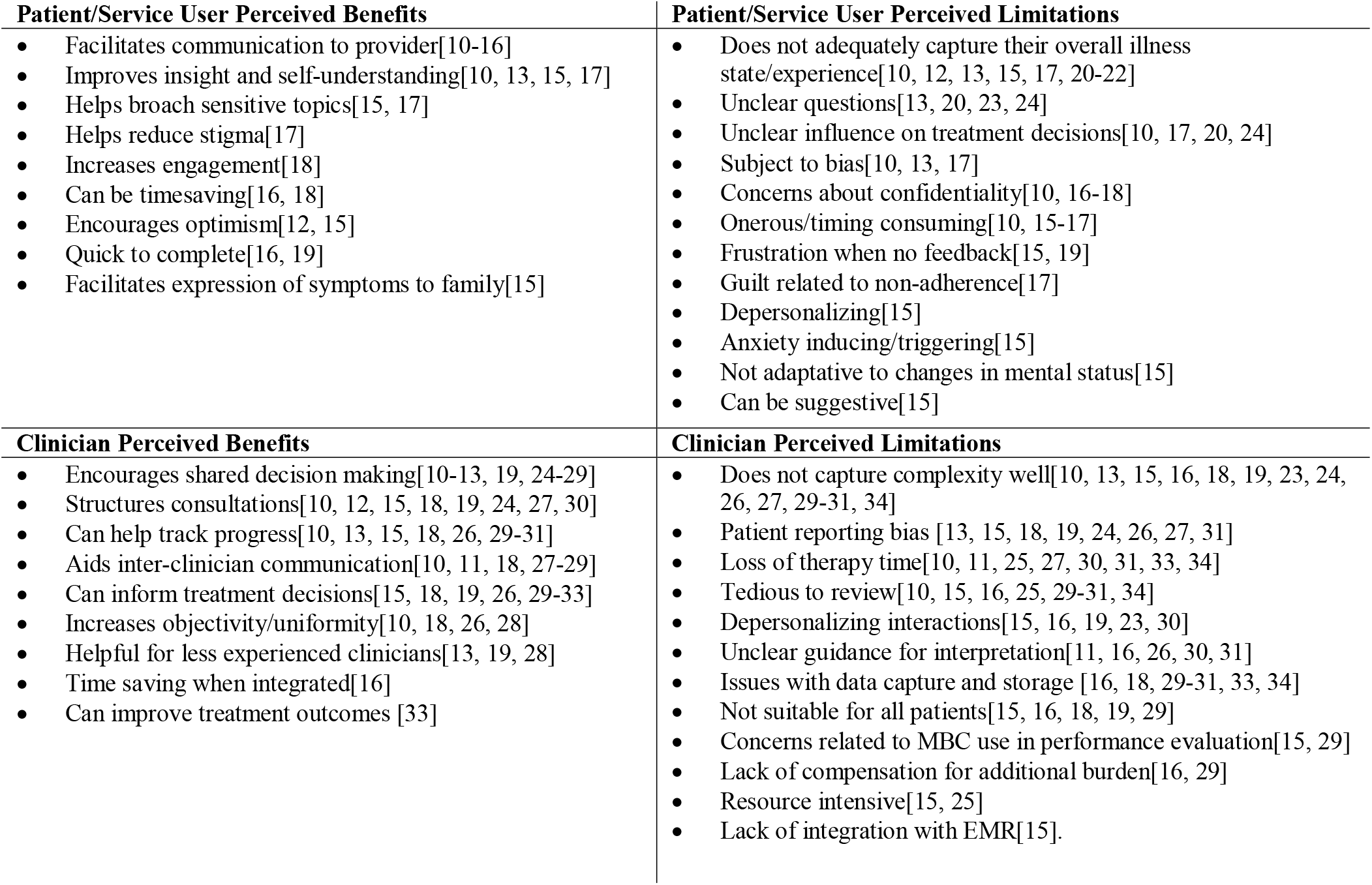

### 3.5 Patient attitudes towards benefits of MBC

With respect to patient reported benefits, the two most common findings were that patients found MBC to be helpful with respect to: 1) allowing another avenue for expressing their experience to providers,[10–13, 15, 17, 19] and 2) improving their own insight into their mental health issues.[10–13, 15, 17] One study reported that approximately 70% of patients found questionnaires to be “quite a bit helpful” in informing health care providers of their health status.[11] Patients also felt MBC helped them to better understand manifestations of their mental health condition.[10, 13, 15, 17] This sentiment held true even for some patients undergoing treatment for borderline personality disorder,[15] who reported that they felt their therapist got a better understanding of them as they were able to convey difficult to describe emotions/symptoms through MBC. At least one study reported that patients found that MBC reduced stigma and was helpful in broaching sensitive topics[17] such as communicating to providers concerns about sexual dysfunction, and with loved ones about their illness experience.[15] This same study reported that patients felt that MBC could be timesaving, for example if used in a waiting room situation, and could help focus assessment around pertinent and active mental health symptoms.[17] Two studies reported patient claims that MBC provided them a clear, concrete reason for optimism around their own improvement.[12, 15] Patients also generally found MBC quick and easy to use and interpret.[19] Notably, in contrast to concerns expressed by care providers that older patients might find use of electronic platforms for completion of MBC to be challenging, many older adults and carers were enthusiastic about using electronic platforms and reported using computers on a regular basis.[18] Patients overall felt that MBC enabled them to become more active participants in their care provided feedback was shared with them.

### 3.6 Patient reported limitations of MBC

The most reported limitation was that the scales/questionnaires utilized did not adequately describe or reflect their overall clinical state or experience.[10, 12, 13, 15, 17, 20–22] For example, one of the studies reported that less than a quarter of patient respondents indicated satisfaction with the accuracy of their QIDS questionnaire,[22] while others noted concerns with the wording of certain questionnaires that can negatively interfere with interpretation.[13, 20, 21, 24] The next most frequent concern from patients was related to their uncertainty in how their questionnaire responses and scores would affect their overall treatment.[10, 17, 20, 24] Related to these concerns, Cuperfain et al. (2021) and Wolpert et al. (2014), noted that some patients admitted to altering or obfuscating their true answers to certain questions in order to avoid the risk of changes being made to their care in response. This also relates to patient insecurity about admitting their own condition to themselves and others, as well as concerns around the confidentiality of who can see their scores or even use them to take medical action. Other patient identified limitations include tediousness of repeat questionnaire completion,[10, 15, 17] concerns about straining patient-provider relations,[17] and frustration when no feedback is provided following scale completion.[15, 19] There are also expressed concerns that completion of MBC-related questionnaires may be upsetting for some via unnecessary triggering of latent memories of negative events/thoughts/feelings which in turn may invoke emotional distress in the absence of a therapist.[15] The same study also noted concerns that targeted questionnaires may prove suggestive for patients with unstable personality structures, and recorded dissatisfaction from patients around completing the same non-adaptive scale throughout their treatment and improvement process.[15]

### 3.7 Clinician perspectives towards benefits of MBC

Our review also identified several perceived benefits of MBC reported by clinicians. These benefits include the belief that MBC helps increase patient involvement in shared decision making surrounding modification of treatment,[10–13, 19, 24–28] that MBC aids in tracking treatment response,[10, 13, 15, 18, 26, 30, 31] and its potential for improving efficiency of focused consultative interviews,[10, 12, 18, 19, 24, 27, 30] provided that self-administered scales and questionnaires are completed prior to the appointment. Many clinicians also found that MBC can be helpful with respect to facilitating efficient inter-clinician communication by quantifying disease severity in a standardized manner (e.g. a Yale-Brown Obsessive-Compulsive Scale score above 24 indicating severe obsessive-compulsive disorder).[10, 11, 18, 27, 28] Beyond tracking treatment effects, some clinicians believed that such scales help increase objectivity and inform treatment decisions made by providers (e.g., distinguishing between mild versus moderate disease in the context of limited functional impairment). This can be particularly helpful for less experienced clinicians such as junior residents,[13, 19, 28] or general practitioners who may not be as familiar with diagnostic criteria and are deciding whether to refer for specialist care or not. Clinicians spoke positively of the potential for the integration of MBC into clinical documentation as a time-saving measure.[19]

### 3.8 Clinician perspectives towards limitations of MBC

These benefits are balanced by reporting of several common limitations of MBC. The most frequently reported limitation included concerns that scales used for MBC did not adequately capture complexity (a view shared by 12 studies).[13, 15, 18, 19, 21, 23, 24, 26, 27, 30, 31, 34] Specifically, clinicians felt that many scales do poorly in patients presenting with atypical presentations or when patients have multiple comorbid conditions that interact. The second most reported limitation was concerns related to biases in patient reporting (reported in 9 studies) due to both conscious and unconscious influences such as the desire to please the clinician and show progress.[13, 15, 18, 19, 21, 24, 26, 27, 31] Some clinicians also felt that incorporating MBC into routine care would be too time consuming and would result in loss of valuable session time,[10, 11, 15, 25, 27, 30, 31, 34] especially when scales are particularly tedious or difficult to review due to lack of integration into the EMR system.[10, 21, 25, 30, 31, 34] Other clinicians expressed worries that MBC can result in depersonalization of interactions and thus negatively impact the therapeutic alliance.[15, 19, 21, 23, 30] There were also expressed concerns about unclear guidance for how to interpret MBC results related to limited training on MBC,[11, 26, 30, 31] as well as frustration related to logistical issues of data capture and storage.[18, 30, 31, 34] Data use troubled some physicians, as far as the potential for its use in performance evaluation.[15, 29] Notably, while one large study of 922 clinicians reported that concern around data use was the most strongly endorsed barrier to using MBC, these concerns do not correlate with a decrease in use of MBC – rather the greatest decrease in MBC use was perceived low clinical utility followed by administrative burden.[29] Some clinicians also perceived MBC as an increase in workload and administrative burden, for which they felt they should be compensated.[19, 29] Lastly, there were concerns that MBC is not suitable for all patients – for example those with cognitive difficulties, patients who are illiterate, patients with poor insight, and those not fluent in English.[18–20]

## 4. Discussion

The implementation of Measurement-Based Care (MBC) in mental health services has garnered significant attention of recent years. Patients and clinicians alike acknowledge the potential of MBC to transform the therapeutic landscape, albeit with reservations and highlighted limitations that warrant careful consideration.

From the patient’s perspective, the benefits of MBC are notably centered around enhancing communication with healthcare providers and fostering a deeper understanding of their own mental health conditions. The ability for patients to express their experiences through standardized scales and questionnaires, as reported in the studies reviewed, not only facilitates a more open dialogue with clinicians but also empowers patients by granting them insight into the nuances of their mental health.[10, 13, 15, 17] This dual advantage underscores the potential of MBC to bridge the communication gap between patients and providers, thereby promoting a more collaborative approach to care. The efficiency and ease of utilizing MBC, including via electronic platforms, facilitates that communication and collaboration in a time-saving manner.[16, 18, 19] Moreover, the positive reception of MBC among older adults, and their carers (particularly in the context of electronic platform usage), challenges preconceived notions about the technological adeptness of this demographic.[18] This finding suggests a broader acceptability and potential for the integration of MBC across various patient populations, thereby enhancing its applicability in diverse clinical settings. However, the discussion of benefits would be incomplete without addressing the limitations highlighted by patients. The primary concern revolves around the adequacy of MBC tools in capturing the full spectrum of the patient’s clinical state.[10, 12, 13, 17, 20–22] Some worry about the suggestiveness of a scale that is targeted to a single condition,[15] while others doubt the accuracy of a scale that does not adapt alongside their condition.[15] This gap between patient experiences and the quantitative assessments offered by MBC scales raises questions about the comprehensiveness and sensitivity of assessment tools captured in the articles reviewed. Furthermore, the apprehension regarding the impact of questionnaire responses on treatment decisions points to a need for greater transparency and communication about the role of MBC in clinical decision-making.[10, 15, 18, 22]

Clinician perspectives offer a professional counterpoint to patient views, emphasizing the utility of MBC in enhancing patient involvement, tracking treatment response, and streamlining clinical communication.[10–13, 19, 24–28] There was also relative consensus among clinicians regarding the value of MBC in objectifying mental health assessments and informing treatment adjustments. This could reflect a broader trend towards evidence-based practices in healthcare. Yet, clinicians are also cognizant of the limitations, particularly concerning the complexity of mental health conditions that may not be fully captured by standardized scales.[13, 16, 17, 19, 21, 22, 24, 25, 27–29] Additionally, concerns regarding patient reporting biases and the potential for MBC to encroach on the therapeutic alliance further underscore the nuanced challenges inherent in integrating MBC into routine care.[13, 16, 17, 19, 22, 24, 25, 28] Concerns around a lack of compensation for what they foresee to be additional burden,[16, 29] and fear of the potential for MBC data to become a metric for performance evaluation,[15, 29] help illuminate initial clinician apprehension toward MBC implementation.

### 4.1 Moving forward

In synthesizing these perspectives, it becomes evident that while MBC holds considerable promise in enhancing mental health care, its successful implementation hinges on addressing the outlined limitations. We believe this can be achieved through three guiding principles for MBC implementation – standardization, transparency, and simplification. This involves the development of more nuanced and comprehensive assessment tools, clinician training on how to effectively use MBC, development of guidelines regarding the use and interpretation of assessment tools, clear communication regarding the role of MBC in treatment planning, routine patient feedback, and use of strategies to maintain the therapeutic alliance in the context of MBC. Furthermore, the integration of MBC into clinical practice must be approached with sensitivity to the diverse needs and concerns of both patients and clinicians, ensuring that the move towards standardized assessments does not eclipse the individualized nature of mental health care, while at the same not adding to the growing administrative burden experienced by clinicians. This can be theoretically achieved through automation and integration into EMR systems to facilitate data collection and ease of tracking.

### 4.2 Strengths and Limitations

Strengths of this review include the large number of studies screened, the quality appraisal, the inclusion of mostly moderate to high quality studies, studies that range from children and youth[24, 25] to older adults,[18] and the comparison of similarities and differences in perceptions of MBC between patients and clinicians. Limitations include the exclusion of grey literature, of non-English articles and the large heterogeneity between studies that restricted analysis to a narrative format. Another limitation is that the studies included commonly excluded patients with communication problems or significant cognitive impairment – patients who may understandably have the most difficulty completing MBC independently.

## 5. Conclusions & Clinical Implications

In conclusion, the findings of this study highlight a complex landscape where the potential of MBC to revolutionize mental health care is tempered by significant challenges. While champions of MBC attest that it holds significant promise with respect to enhancing mental health treatment by enabling better patient-centered care through increased patient engagement, increasing efficiency of clinical encounters, and supporting clinical decision making through monitoring of treatment progress, public opinions about it remain mixed. To realize the full potential of MBC, the path forward lies in a collaborative effort to refine MBC tools and practices, ensuring that they enhance, rather than detract from, the quality and personalization of mental health services. It will also require efforts improve training and increase integration of such tools into current healthcare systems.

## Supporting information

supplemental index 1

supplemental index 2

## Data Availability

All data produced in the present study are available upon reasonable request to the authors.

## Data availability statement

Data sharing not applicable as no datasets generated and/or analysed for this study. All data relevant to the study are included in the article or uploaded as online supplemental information. All data underlying the results are available as part of the article and no additional source data are required.

## Ethics statements

Patient consent for publication - Not required.

Ethics approval - No ethical approval was needed because data from previous published studies are being used.

## Acknowledgments/Disclosures

No conflicts of interest to disclose.

## Notes

### Competing Interest Statement

The authors have declared no competing interest.

### Clinical Protocols

https://www.crd.york.ac.uk/PROSPERO/view/CRD420250651562

### Funding Statement

This study did not receive any funding.

