## supplemental index 2 for "Quantifying Care, Qualifying Experiences: A Systematic Review of Measurement-Based Care in Psychiatry from Patient and Provider Perspectives"

**Supplemental Index 1**

**Prospero Search Strategy**
The search strategy was developed for the electronic databases to identify journal articles. The following databases were searched for related studies: Ovid MEDLINE (1946 to present), EMBASE (Classic EMBASE + EMBASE ((1947 to present)), EBM Reviews – Cochrane Central Register of Controlled Trials (1991 to January 2024), APA PsychINFO (1806 to June 2024), and CINAHL (via EBSCO ((1981 to current)). The search strategy was designed to comprehensively capture literature related to measurement-based care in mental health, with a focus on patient and physician perspectives. Specifically, the search combined three main concepts: 1) mental health including MeSH terms and keywords for psychiatry, mental disorders, anxiety disorders, bipolar disorder and mood disorders; 2) measurement based care, encompassing terms related to routine outcome monitoring, patient-reported measures, and feedback-informed treatment in mental health settings; and 3) patient and physician perspectives, incorporating MeSH terms and keywords to capture viewpoints and attitudes of both patients and healthcare providers. These concepts were combined using Boolean operators, with results limited to English language and human studies. Notably while, searches in the electronic databases were conducted from database inception until January 2024. The search was later limited to studies reported in English, involving human subjects. While initially there was no limit on publication year, only papers published during or after 2000 were included.

Platforms and Databases Searched:

| **Platform** | **Database** | **Coverage Dates** | **# Rec.** | **Search Date** | **Notes** |
| --- | --- | --- | --- | --- | --- |
| OvidSP | MEDLINE ® and Epub Ahead of Print, In-Process, In-Data-Review & Other Non-Indexed Citations, Daily and Versions ® | 1946 – Current | 230 | 20 Jan 2024 | Limits applied: Human and English language |
| OvidSP | Classic Embase + Embase | 1947 – 2024 | 646 | 20 Jan 2024 | Limits applied: Human and English language |
| Ovid SP | EBM Reviews – Cochrane Central Register of Controlled Trials | 1991 –2024 Week 4 | 20 | 20 Jan 2024 | Limits applied: English language |
| Ovid SP | APA PsycINFO | 1806 – Jan Week 4 2024 | 168 | 20 Jan 2024 | Limits applied: Human and English language |
| EBSCO | CINAHL | 1981 - current | 267 | 20 Jan 2024 | Limits applied: Human and English language |

Total # of records prior to duplicate removal: 1643

Total # of records after removing duplicates: 1432

Original Searches:

**MEDLINE® and Epub Ahead of Print, In-Process, In-Data-Review & Other Non-Indexed Citations, Daily and Versions®**

Database: Ovid MEDLINE(R) and Epub Ahead of Print, In-Process, In-Data-Review & Other Non-Indexed Citations, Daily and Versions(R) <1946 to Jan 20, 2024> Search Strategy:

--------------------------------------------------------------------------------

1	exp Child Psychiatry/ or exp Psychiatry/ or exp Adolescent Psychiatry/ (107836)

2	exp mental health/ or exp mental disorders/ or exp anxiety disorders/ or exp "bipolar and related disorders"/ or exp mood disorders/ (1379273)

3	exp Depression/ (136683)

4	exp Depressive Disorder/ (116200)

5	(psychiatr* or child psych* or adolescent psych*).mp. (383726)

6	(mental health or mental disorder* or mental health disorder* or mental health issue* or anxiety or anxiety disorder* or bipolar* or mood disorder*).mp. (699435)

7	(depression or depressive disorder* or major depressive disorder*).mp. (484664)

8	or/1-7 [Mental Health cluster] (2008860)

9	(measurement based care or measurement-based care).mp. (329)

10	((mental health or psych*) adj3 questionnaire).ti,hw,kf. (678)

11	feedback informed treatment.mp. (17)

12	(routine* adj2 monitor* adj2 (client* or progress*)).mp. (21)

13	"outcome* monitoring and feedback".mp. (19)

14	patient* focused research.mp. (48)

15	patient* level feedback.mp. (8)

16	patient* reported outcome* measure*.mp. (17724)

17	routine outcome* monitoring.mp. (272)

18	patient-reported outcome* monitoring.mp. (23)

19	or/9-18 [Measurement-based care cluster] (19031)

20	exp Patients/ or exp Physicians/ (231325)

21	((perspective* or view* or opinion* or attitude*) adj1 (patient* or physician*)).mp. (25685)

22	20 or 21 (252442)

23	8 and 19 and 22 (251)

24	limit 23 to (english language and humans) (230)

***************************

**Embase**

Database: Embase Classic+Embase <1947 to 2024 Week 4> Search Strategy:

--------------------------------------------------------------------------------

1	exp child psychiatry/ or exp psychiatry/ (156149)

2	exp mental health/ (193232)

3	exp mental disease/ (2542546)

4	exp anxiety disorder/ or exp anxiety/ (492419)

5	exp major depression/ or exp depression/ or exp bipolar depression/ or exp long term depression/ (546654)

6	(psychiatr* or child psych* or adolescent psych*).mp. (493435)

7	(mental health or mental disorder* or mental disease* or mental health disorder* or mental health issue* or anxiety or anxiety disorder* or bipolar* or mood disorder*).mp. (1061515)

8	(depression or depressive disorder* or major depression or major depressive disorder* or long term depression).mp. (783223)

9	or/1-8 [Mental health cluster] (3192337)

10	(measurement based care or measurement-based care).mp. (411)

11	((mental health or psych*) adj3 questionnaire).ti,hw,kw. (1780)

12	feedback informed treatment.mp. (20)

13	(routine* adj2 monitor* adj2 (client* or progress*)).mp. (28)

14	"outcome* monitoring and feedback".mp. (20)

15	patient* focused research.mp. (76)

16	patient* level feedback.mp. (11)

17	patient* reported outcome.mp. (44238)

18	routine outcome* monitoring.mp. (332)

19	patient-reported outcome* monitoring.mp. (33)

20	or/10-19 [Measurement-based care cluster] (46772)

21	exp physician attitude/ (55917)

22	((perspective* or opinion* or view* or attitude*) adj2 (patient* or physician*)).mp. (170734)

23	21 or 22 (170734)

24	9 and 20 and 23 (670)

25	limit 24 to (human and english language) (646)

***************************

**EBM Reviews – Cochrane Central Register of Controlled Trials\**

Database: EBM Reviews - Cochrane Central Register of Controlled Trials <Jan 2024> Search Strategy:

--------------------------------------------------------------------------------

1	exp Child Psychiatry/ or exp Psychiatry/ or exp Adolescent Psychiatry/ (502)

2	exp mental health/ or exp mental disorders/ or exp anxiety disorders/ or exp "bipolar and related disorders"/ or exp mood disorders/ (76078)

3	exp Depression/ (13443)

4	exp Depressive Disorder/ (12991)

5	(psychiatr* or child psych* or adolescent psych*).mp. (34161)

6	(mental health or mental disorder* or mental health disorder* or mental health issue* or anxiety or anxiety disorder* or bipolar* or mood disorder*).mp. (94650)

7	(depression or depressive disorder* or major depressive disorder*).mp. (89769)

8	or/1-7 [Mental Health cluster] (193880)

9	(measurement based care or measurement-based care).mp. (68)

10	[((mental health or psych*) adj3 questionnaire).ti,hw,kf.] (0)

11	feedback informed treatment.mp. (13)

12	(routine* adj2 monitor* adj2 (client* or progress*)).mp. (4)

13	"outcome* monitoring and feedback".mp. (6)

14	patient* focused research.mp. (6)

15	patient* level feedback.mp. (5)

16	patient* reported outcome* measure*.mp. (2875)

17	routine outcome* monitoring.mp. (40)

18	patient-reported outcome* monitoring.mp. (5)

19	or/9-18 [Measurement-based care cluster] (3014)

20	exp Patients/ or exp Physicians/ (8002)

21	((perspective* or view* or opinion* or attitude*) adj1 (patient* or physician*)).mp. (8361)

22	20 or 21 (15804)

23	8 and 19 and 22 (25)

24	limit 23 to (english language and humans) [Limit not valid; records were retained] (20)

***************************

**APA PsycINFO**

Database: APA PsycInfo <1806 to Jan Week 4 2024> Search Strategy:

--------------------------------------------------------------------------------

1	exp Psychiatry/ or exp Adolescent Psychiatry/ or exp Child Psychiatry/ (54250)

2	exp mental disorders/ or exp mental health/ (972627)

3	exp Anxiety Disorders/ or exp Anxiety/ (128928)

4	exp "Depression (Emotion)"/ or exp Major Depression/ (169682)

5	(psychiatr* or child psych* or adolescent psych*).mp. (384984)

6	(mental health or mental disorder* or mental health disorder* or mental health issue* or anxiety or anxiety disorder* or bipolar* or mood disorder*).mp. (646136)

7	(depression or depressive disorder* or major depressive disorder*).mp. (369992)

8	or/1-7 [Mental Health Cluster] (1442915)

9	(measurement based care or measurement-based care).mp. (268)

10	((psych* or mental health) adj3 questionnaire).ti,ab,hw. (4488)

11	feedback informed treatment.mp. (56)

12	(routine* adj2 monitor* adj2 (client* or progress*)).mp. (17)

13	"outcome* monitoring and feedback".mp. (19)

14	patient* focused research.mp. (81)

15	patient* level feedback.mp. (2)

16	patient* reported outcome*.mp. (4001)

17	routine outcome* monitoring.mp. (336)

18	patient-reported outcome* monitoring.mp. (4)

19	or/9-18 [Measurement-based care cluster] (9171)

20	exp Health Personnel Attitudes/ (25439)

21	((perspective* or view* or opinion* or attitude*) adj2 (patient* or physician*)).mp. (12782)

22	20 or 21 (36123)

23	8 and 19 and 22 (198)

24	limit 23 to (human and english language) (168)

***************************

**CINAHL**

| **Search ID#** | **Search Terms** | **Search Options** | **Last Run Via** | **Results** |
| --- | --- | --- | --- | --- |
| S26 | S9 AND S20 AND S24 | Limiters - English Language; Human Expanders - Apply equivalent subjects Search modes - Boolean/Phrase | Interface - EBSCOhost Research Databases Search Screen - Advanced Search Database - CINAHL | 267 |
| S25 | S9 AND S20 AND S24 | Expanders - Apply equivalent subjects Search modes - Boolean/Phrase | Interface - EBSCOhost Research Databases Search Screen - Advanced Search Database - CINAHL | 331 |
| S24 | S21 OR S22 OR S23 | Expanders - Apply equivalent subjects Search modes - Boolean/Phrase | Interface - EBSCOhost Research Databases Search Screen - Advanced Search Database - CINAHL | 89,454 |
| S23 | TX ((perspective* or opinion* or view* or attitude*) N2 (patient* or physician*)) | Expanders - Apply equivalent subjects Search modes - Boolean/Phrase | Interface - EBSCOhost Research Databases Search Screen - Advanced Search Database - CINAHL | 89,454 |
| S22 | (MH "Physician Attitudes") | Expanders - Apply equivalent subjects Search modes - Boolean/Phrase | Interface - EBSCOhost Research Databases Search Screen - Advanced Search Database - CINAHL | 16,468 |
| S21 | (MH "Patient Attitudes") | Expanders - Apply equivalent subjects Search modes - Boolean/Phrase | Interface - EBSCOhost Research Databases Search Screen - Advanced Search Database - CINAHL | 53,260 |
| S20 | S10 OR S11 OR S12 OR S13 OR S14 OR S15 OR S16 OR S17 OR S18 OR S19 | Expanders - Apply equivalent subjects Search modes - Boolean/Phrase | Interface - EBSCOhost Research Databases Search Screen - Advanced Search Database - CINAHL | 19,443 |
| S19 | TX patient-reported outcome* monitoring | Expanders - Apply equivalent subjects Search modes - Boolean/Phrase | Interface - EBSCOhost Research Databases Search Screen - Advanced Search Database - CINAHL | 8 |
| S18 | TX routine outcome* monitoring | Expanders - Apply equivalent subjects Search modes - Boolean/Phrase | Interface - EBSCOhost Research Databases Search Screen - Advanced Search Database - CINAHL | 122 |
| S17 | TX patient* reported outcome | Expanders - Apply equivalent subjects Search modes - Boolean/Phrase | Interface - EBSCOhost Research Databases Search Screen - Advanced Search Database - CINAHL | 15,232 |
| S16 | TX patient* level feedback | Expanders - Apply equivalent subjects Search modes - Boolean/Phrase | Interface - EBSCOhost Research Databases Search Screen - Advanced Search Database - CINAHL | 6 |
| S15 | TX patient* focused research | Expanders - Apply equivalent subjects Search modes - Boolean/Phrase | Interface - EBSCOhost Research Databases Search Screen - Advanced Search Database - CINAHL | 19 |
| S14 | TX "outcome* monitoring and feedback" | Expanders - Apply equivalent subjects Search modes - Boolean/Phrase | Interface - EBSCOhost Research Databases Search Screen - Advanced Search Database - CINAHL | 8 |
| S13 | TX (routine* N2 monitor* N2 (client* or progess*)) | Expanders - Apply equivalent subjects Search modes - Boolean/Phrase | Interface - EBSCOhost Research Databases Search Screen - Advanced Search Database - CINAHL | 4 |
| S12 | TX feedback informed treatment | Expanders - Apply equivalent subjects Search modes - Boolean/Phrase | Interface - EBSCOhost Research Databases Search Screen - Advanced Search Database - CINAHL | 8 |
| S11 | TX ((mental health or psych*) N2 questionnaire) | Expanders - Apply equivalent subjects Search modes - Boolean/Phrase | Interface - EBSCOhost Research Databases Search Screen - Advanced Search Database - CINAHL | 3,961 |
| S10 | TX (measurement based care or measurement-based care) | Expanders - Apply equivalent subjects Search modes - Boolean/Phrase | Interface - EBSCOhost Research Databases Search Screen - Advanced Search Database - CINAHL | 150 |
| S9 | S1 OR S2 OR S3 OR S4 OR S5 OR S6 OR S7 OR S8 | Expanders - Apply equivalent subjects Search modes - Boolean/Phrase | Interface - EBSCOhost Research Databases Search Screen - Advanced Search Database - CINAHL | 1,011,015 |
| S8 | TX (depression or depressive disorder* or major depressive disorder*) | Expanders - Apply equivalent subjects Search modes - Boolean/Phrase | Interface - EBSCOhost Research Databases Search Screen - Advanced Search Database - CINAHL | 203,229 |
| S7 | TX (mental health or mental disorder* or mental health disorder* or mental health issue* or anxiety or anxiety disorder* or bipolar* or mood disorder*) | Expanders - Apply equivalent subjects Search modes - Boolean/Phrase | Interface - EBSCOhost Research Databases Search Screen - Advanced Search Database - CINAHL | 405,211 |
| S6 | TX (psychiatr* or child psych* or adolescent psych*) | Expanders - Apply equivalent subjects Search modes - Boolean/Phrase | Interface - EBSCOhost Research Databases Search Screen - Advanced Search Database - CINAHL | 429,901 |
| S5 | (MH "Depression+") OR (MH "Bipolar Disorder+") | Expanders - Apply equivalent subjects Search modes - Boolean/Phrase | Interface - EBSCOhost Research Databases Search Screen - Advanced Search Database - CINAHL | 133,775 |
| S4 | (MH "Mental Disorders+") OR (MH "Mental Disorders, Chronic") | Expanders - Apply equivalent subjects Search modes - Boolean/Phrase | Interface - EBSCOhost Research Databases Search Screen - Advanced Search Database - CINAHL | 609,901 |
| S3 | (MH "Anxiety+") OR (MH "Anxiety Disorders+") | Expanders - Apply equivalent subjects Search modes - Boolean/Phrase | Interface - EBSCOhost Research Databases Search Screen - Advanced Search Database - CINAHL | 95,178 |
| S2 | (MH "Mental Health") | Expanders - Apply equivalent subjects Search modes - Boolean/Phrase | Interface - EBSCOhost Research Databases Search Screen - Advanced Search Database - CINAHL | 45,877 |
| S1 | (MH "Psychiatry+") OR (MH "Child Psychiatry") OR (MH "Adolescent Psychiatry") | Expanders - Apply equivalent subjects Search modes - Boolean/Phrase | Interface - EBSCOhost Research Databases Search Screen - Advanced Search Database - CINAHL | 18,057 |
