## supplementary table S1 for "Quantifying Care, Qualifying Experiences: A Systematic Review of Measurement-Based Care in Psychiatry from Patient and Provider Perspectives"

| **Author &**  **Year** | **Study Design** | **Quality Appraisal** | **Psych Scales Used** | **Participants** | | **Patient Population** | **Clinical Setting** | **Endorsed Clinician Perspectives** | **Endorsed Patient Perspectives** |
| --- | --- | --- | --- | --- | --- | --- | --- | --- | --- |
|  |  |  |  | **# clinicians** | **# patients** |  |  |  |  |
| Bailey et al. 2023.[16] | Mixed Methods | Medium | N/A | 17 | 40 | Adult | Outpatient | - Time savings when integrated - Does not capture complexity well  -Tedious to review - Depersonalizing interactions - Unclear guidance for interpretation - Concerns with data capture and storage - Not suitable for all patients - Lack of compensation for additional burden. | - Facilitates communication with provider - Can be time saving - Quick to complete  - Concerns about confidentiality - Can be onerous to complete - Frustration when no feedback |
| Brand et al. 2022.[15] | Qualitative | High | BPDSI-IV | 10 | 10 | Adult (borderline) | Outpatient | - Structure consultations - Helpful for tracking progress - Can inform treatment-decisions - Does not capture complexity well - Subject to patient reporting bias -Tedious to review - Depersonalizing interactions - Not suitable for all patients - Concerns related to MBC use in performance evaluation  - Resource intensive - Lack of integration with EMR | - Improves insight and self-understanding - Facilitates communication with provider  - Helps broach sensitive topics  - Encourages optimism  - Facilitates expression of symptoms to family  - Does not adequately overall capture illness experience  - Can be onerous to complete - Frustration when no feedback  - Depersonalizing  - Anxiety Inducing  - Not adaptative to changes in MSE  - Can be suggestive. |
| Burr et al. 2017.[31] | Mixed Methods | Low | PHQ-9, Somatic, Anxiety  Columbia Suicide Severity Rating Scale  Relationship Questionnaire  Difficulties in Emotional Regulation Scale  Acceptance and Action Questionnaire  WHO Well Being Index  WHODAS 2.0  Stressful Life Events Inventory  Menninger Quality of Care Measure | 70 | N/A | Adult  (mixed disorders) | Inpatient | - Helpful for tracking progress - Can inform treatment decisions - Does not capture complexity well - Subject to patient reporting bias  - Loss of therapy time - Tedious to review - Unclear guidance for interpretation - Concerns with data capture and storage |  |
| Cuperfain et al. 2021.[10] | Qualitative | Medium | Clinical Global Impression (CGI) Brief Psychiatric Rating Scale (BPRS)  Quality of Life and Enjoyment Satisfaction Questionnaire (QLES-Q) Abnormal Involuntary Movement Scale (AIMS) | 14 | 22 | Young Adults (psychotic disorders) | Outpatient | - Encourages shared decision-making - Structures consultations - Helpful for tracking progress - Aids inter-clinician-communication - Increases objectivity  - Does not capture complexity well  - Loss of therapy time - Tedious to review | - Improves insight and self-understanding - Facilitates communication with provider - Does not adequately overall capture illness experience  - Subject to bias - Concerns about confidentiality - Can be onerous to complete |
| Egeter et al. 2018.[11] | Quantitative | Medium | Brief Symptom Inventory  WHO Quality of Life Short Form  Beck Depression Inventory II  Hospital Anxiety and Depression Scale  University of Rhode Island Change Assessment Scale  Brief Illness Perception Questionnaire  Sleep Quality and Pain Intensity | 23 | 164 | Adult (mixed disorders) | Inpatient | - Encourages shared decision-making - Aids inter-clinician-communication - Loss of therapy time - Unclear guidance for interpretation | - Facilitates communication with provider |
| Greenberg et al. 2021.[25] | Quantitative | Medium | Paediatric Quality of Life Inventory (PedsQL) Canadian Occupational Performance Measure  Community Assessment of Psychic Experience (CAPE)  Paediatric Outcomes Data Collection Instrument | 113 | N/A | Children-Young Adults (ages 8 to 21) | Outpatient | - Encourages shared decision-making - Can improve treatment outcomes - Loss of therapy time - Tedious to review - Resource intensive |  |
| Hawkins et al. 2023.[33] | Quantitative | Medium | PHQ9 | 23 | N/A | Adults (mood disorders) | Outpatient | - Can inform treatment decisions  - Loss of therapy time - Concerns with data capture and storage |  |
| Keepers et al. 2023.[29] | Quantitative | High | N/A | 922 |  | Adults | Outpatient | - Encourages shared decision-making - Helpful for tracking progress - Aids inter-clinician-communication - Can inform treatment decisions - Does not capture complexity well - Tedious to review - Concerns with data capture and storage - Not suitable for all patients - Concerns related to data use towards clinician performance evaluation - Lack of compensation for additional burden |  |
| Kendrick et al. 2017.[19] | Mixed methods | Medium | PHQ9 Distress Thermometer Analogue Scale | 13 | 14 | Adult (mood disorders) | Outpatient | - Encourages shared decision-making - Structures consultations - Can inform treatment decisions - Helpful for less experienced clinicians  - Does not capture complexity well - Patient reporting bias - Depersonalizing interactions - Not suitable for all patients | - Quick to complete - Frustration when no feedback |
| McMullan et al. 2020.[18] | Qualitative | High | N/A | 5 | 23 | Adult (TBI+) | Outpatient | - Structures consultations - Helpful for tracking progress - Aids inter-clinician communication - Increases objectivity - Does not capture complexity well - Patient reporting bias - Not suitable for all patients - Concerns with data capture and storage | - Increases engagement - Can be time saving - Concerns about confidentiality |
| Moltu et al. 2016.[12] | Qualitative | Medium | N/A | 37 | 18 | Geriatric (mixed) | Inpatient & Outpatient | - Encourages shared decision making - Structures consultations | - Encourages optimism - Does not adequately overall capture illness experience |
| Oslin et al. 2019.[26] | Quantitative | Medium | PHQ9  GAD-7  Brief Addiction Monitor  PTSD Checklist (PCL-5) | 230 | N/A | Geriatric (mixed) | Outpatient | - Encourages shared decision making - Helpful for tracking progress - Can inform treatment decisions - Increases objectivity - Does not capture complexity well - Patient reporting bias - Unclear guidance for interpretation |  |
| Perry et al. 2019.[13] | Qualitative | High | Clinical Outcomes in Routine Evaluation (CORE-OM) | 24 | 34 | Adult (psychotic disorders) | Inpatient | - Encourages shared decision making - Helpful for tracking progress - Helpful for less experienced clinicians  - Does not capture complexity well - Patient reporting bias | - Improves insight and self-understanding - Facilitates communication with provider - Does not adequately overall capture illness experience - Subject to bias |
| Robinson et al. 2017.[20] | Qualitative | High | PHQ9 Global Rating of Change | N/A | 29 | Adult (mood disorders) | Outpatient |  | - Does not adequately overall capture illness experience - Unclear influence on treatment decisions - Unclear questions |
| Rye et al. 2019.[23] | Mixed methods | Medium | N/A | 1094 | N/A | Adult | Outpatient | - Does not capture complexity well - Depersonalizing interactions | - Unclear questions |
| Salmond et al. 2020.[21] | Quantitative | Low | Revised Child Anxiety and Depression Scale (RCADS)  Young Person's Clinical Outcome in Routine Evaluation (YP CORE)  Mood's and Feelings Questionnaire (MFQ)  Children's Global Assessment Scale (CGAS)  Health of the nation outcome scales for children and adolescents | N/A | 67 | Young Adults (mixed) | Inpatient | - Does not capture complexity well |  |
| Tauscher et. al. 2021.[27] | Qualitative | High | N/A | 15 | N/A | Adults (substance use disorders) | Outpatient | - Structures consultations - Aids inter-clinician communication - Does not capture complexity well - Patient reporting bias - Loss of therapy time |  |
| Tavabie et al. 2009.[28] | Qualitative | Medium | N/A | 16 | N/A | Adults (Mood Disorders) | Outpatient | - Encourages shared decision making - Aids inter-clinician communication - Increases objectivity  - Helpful for less experienced clinicians |  |
| Van Wert et al. 2020.[30] | Mixed Methods | High | N/A | 180 | N/A | Adult (mixed disorders) | Outpatient | - Structures consultations - Helpful for tracking progress - Can inform treatment decisions - Does not capture complexity well - Loss of therapy time - Tedious to review - Concerns with data capture and storage |  |
| Waldron et al. 2018.[34] | Quantitative | Medium | N/A | 20 | N/A | Child-Young Adult (mixed disorders) | Outpatient | - Does not capture complexity well - Loss of therapy time - Tedious to review - Concerns with data capture and storage |  |
| Wolpert et. al. 2014.[24] | Qualitative | High | N/A | 4 | 6 | Child-Young Adult (mixed disorders) | Outpatient | - Encourages shared decision making  - Structures consultations - Does not capture complexity well - Patient reporting bias | - Unclear influence on treatment decisions |
| Wood et al. 2002.[17] | Qualitative | High | N/A | N/A | 128 | Adult (mixed disorders) | Outpatient |  | - Improves insight and self-understanding - Helps broad sensitive topics - Helps reduce stigma - Does not adequately overall capture illness experience - Unclear influence on treatment decisions - Subject to bias - Can be onerous to complete - Guilt related to non-adherence |
| Zimmerman et al. 2011.[22] | Quantitative | Medium | Quick Inventory of Depressive Symptomology  Remission from Depression Questionnaire | N/A | 102 | Adult (Mood disorders) | Outpatient |  | - Does not adequately overall capture illness experience |
